# Modelling to Predict Hospital Bed Requirements for Covid-19 Patients in California

**DOI:** 10.1101/2020.05.17.20104919

**Authors:** Santanu Basu

## Abstract

A model to predict hospital bed requirements six weeks in advance to treat Covid-19 patients in California is presented. The model also gives prediction for number of cases and deaths during the same period. The model is versatile and can be applied to other countries and regions as well.

## INTRODUCTION

The new coronavirus, SARS-CoV-2 for the disease Covid-19 has spread rapidly through many countries in the world in the first four months of 2020 causing widespread deaths. Mathematical modeling of spreading of infectious diseases is a well-established field [1–2]. There have been a large number of recent efforts in modelling Coronavirus spreading as reported in the literature [3–6]. The Covid-19 cases have put significant strain in the hospital systems around the world. It would be of great benefit to the medical care facilities if the Covid-19 patient load could be known six weeks in advance. This would enable distributing case load among hospitals and to adequately equip the hospitals with staffing, PPE and life support equipment. This is the objective of this paper.

A mathematical model has been recently developed [3] to analyze the Covid-19 case data (time dependent number of cases, deaths and hospitalization) in any country or state. We first present the prediction of Covid-19 cases and hospitalization requirements for the state of California, USA using the model. Next we present the methodology and the model in detail.

## PREDICTION OF HOSPITAL BED REQUIREMENTS IN CALIFORNIA

Covid-19 case data for California was obtained from [7] and the numbers of regular and ICU hospital beds were obtained from the California Health and Human Services websites [8,9]. The first confirmed case of Covid-19 in California was on January 26^th^, 2020. Reliable case numbers became available from March 4^th^, 2020 [7]. Reliable numbers for hospital bed usage since March 29^th^ are available in [8].

The mathematical model used to predict number of cases and hospital loads will be presented in the next section. In this section, we will present the results using the model for the state of California. The model is parametric and transparent so that predictions are based on parameters which are in principle measurable and the predictions can be continuously corrected as more accurate data on parameters become available. Normal curve fitting models which are not based on any real-life parameters tend to have high degree of error in making projections. Also the standard machine learning based and curve-fitting models cannot accurately do what-if experiments, such as what would be the effect of timing of lifting of social distancing measures on the number of cases.

The model has twenty one user-supplied variable parameters, three of which influence the results greatly. These three are (1) the first date of implementation of social distancing protocols since the viral outbreak, (2) the rate at which the rate of transmission decreases by using the government measures and (3) the final rate of transmission in the long run. The California data could be fitted well with March 14^th^ to be the first date (n_1_) of transmission rate decrease, 16 days for exponential time constant for transmission rate decrease (G) and 5% for the final transmission rate (T_f_). The comparison between the model predictions until June 30^th^ and actual data until May 15^th^ on number of cases and deaths are shown in figures 1 and 2 respectively. The figures demonstrate that the model fits the actual data very well.

**Figure 1.**
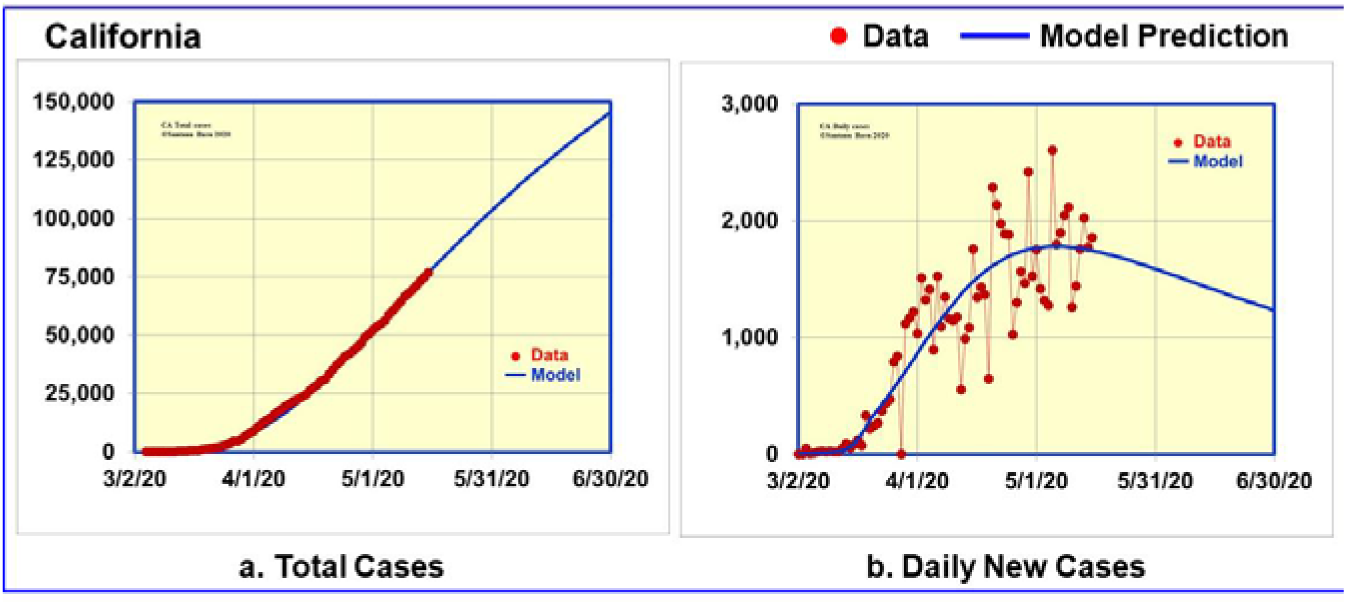
Data and model predictions for Covid-19 cases in California

**Figure 2.**
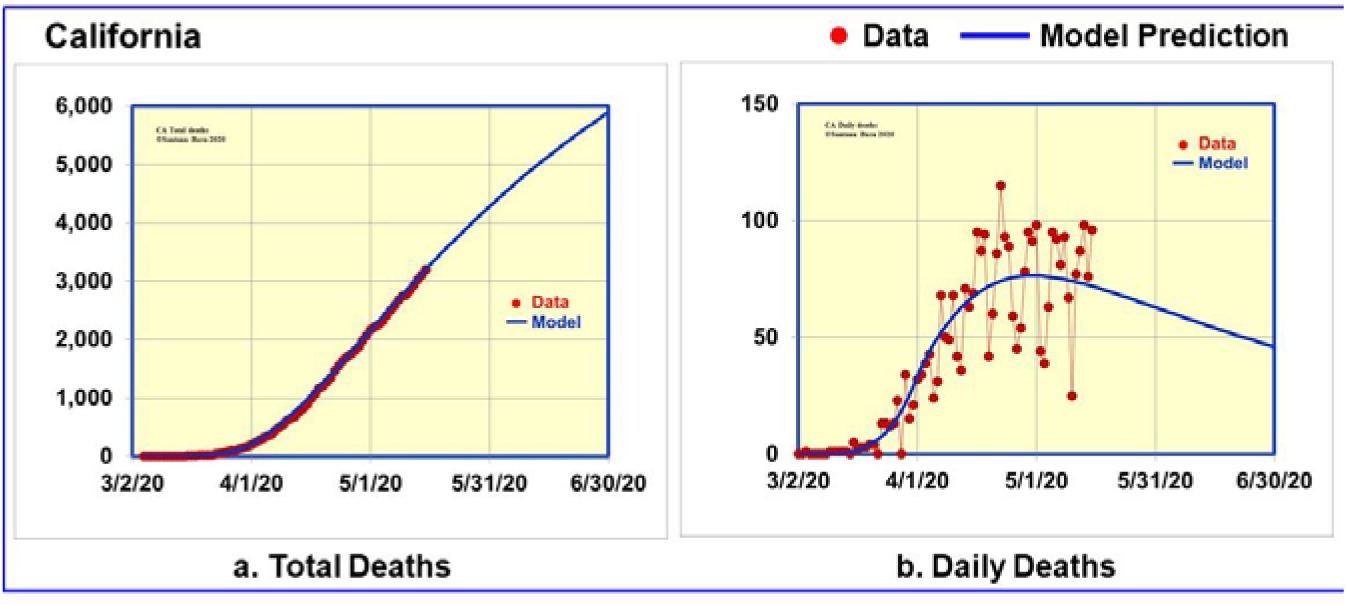
Data and model predictions for Covid-19 deaths in California

Having anchored the model to actual data until May 15^th^, we are in a position to make reliable projections for six weeks in advance. The model predicts the number of cases to rise to 146,000 (range 136,000-151,000) and the number of deaths to rise to 5,900 (range 5600-6,100) on June 30^th^, 2020. The model also predicts the daily number of cases to fall to 1,200 (range 960-1,400) and the number of daily deaths to fall to 45 (range 37-51) on the same date.

**Figure 3.**
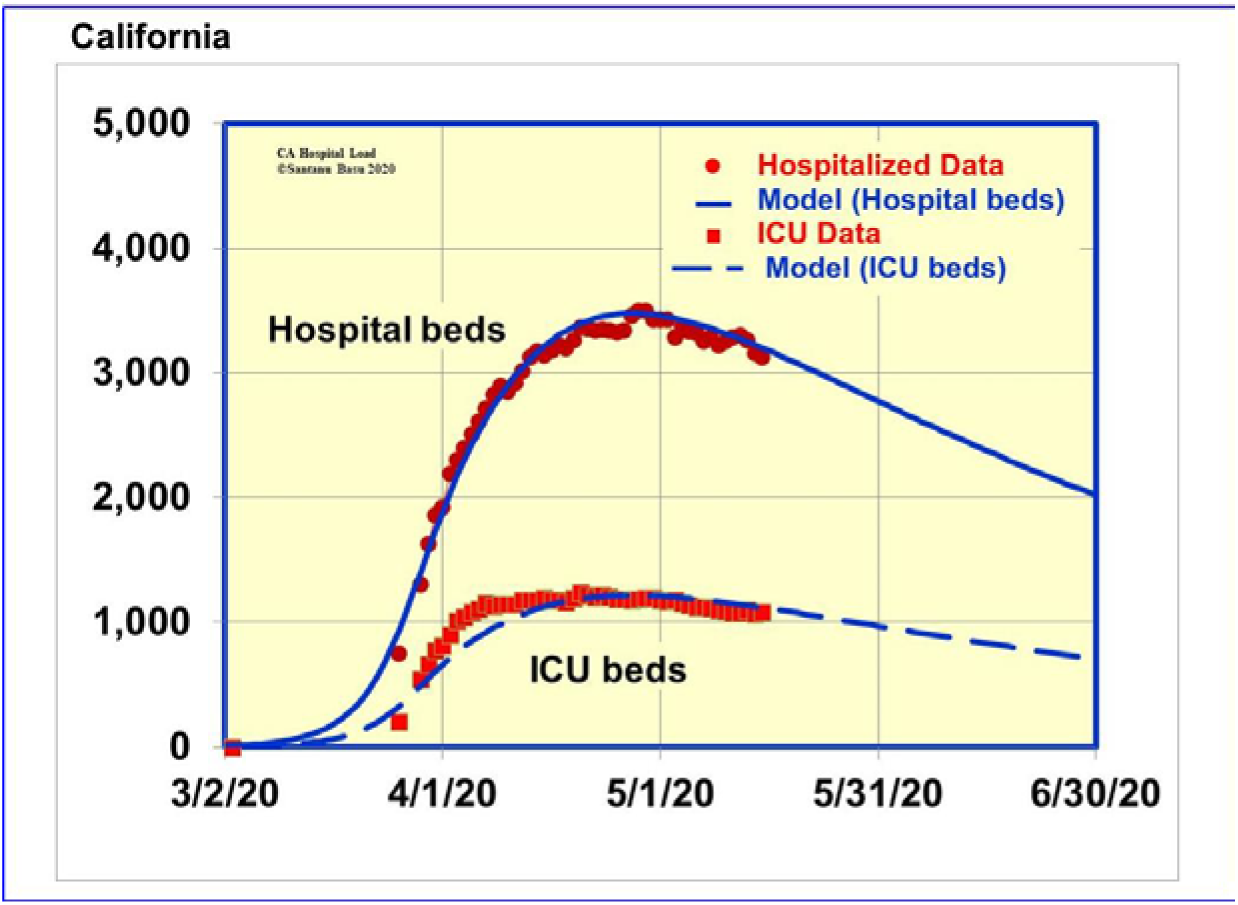
Data and model predictions for Covid-19 hospitalizations in California

Figure 3 shows the comparison between the model predictions until June 30^th^ and actual data until May 15^th^ [8] on number of hospitalizations. We analyzed the data in [8] to determine daily variation of hospitalization rate and the ratio of ICU beds to the total number of hospital beds. The model fits the data very well as shown in figure 3 by assuming 35% of the patients require ICU beds. The model predicts the number of hospitalizations to decrease from 3,248 on May 10^th^ to 2,000 (range 1,600-2,250) on June 30^th^, 2020. The number of ICU beds also will decrease from 1,093 on May 10^th^ to 700 (range 560-790) on June 30^th^, 2020. It is to be noted that the model does not take into account anticipated reopening of businesses in California around the last week of May. It will most likely increase the transmission rate parameter in the model that can be sensed by a perturbation in the trend in the number of cases. The model enables one to make what if exercises such as the effect of relaxing of social distancing.

## METHODOLOGY

In this section we study the propagation of a new infectious disease through an isolated country’s population, P. With some simple modifications, the procedure can be applied to recurring outbreak of the same virus and also to the normal state in which there is population movement among countries.

Figure 4 is a schematic of our model. Let n be the number of days since the onset of virus infection. The entire population of virus infected people is classified in six categories:

I – people in whom the virus is in incubation

A – people who are infected with the virus but who do not exhibit any symptoms

S – non-hospitalized patients who exhibit mild symptoms of the viral infection

H – hospitalized patients who are in acute condition

D – people who have died from the disease

R – people who have recovered from the illness and awaiting to be declared clinically recovered

R^C^ – people who are counted as clinically recovered

**Figure 4.**
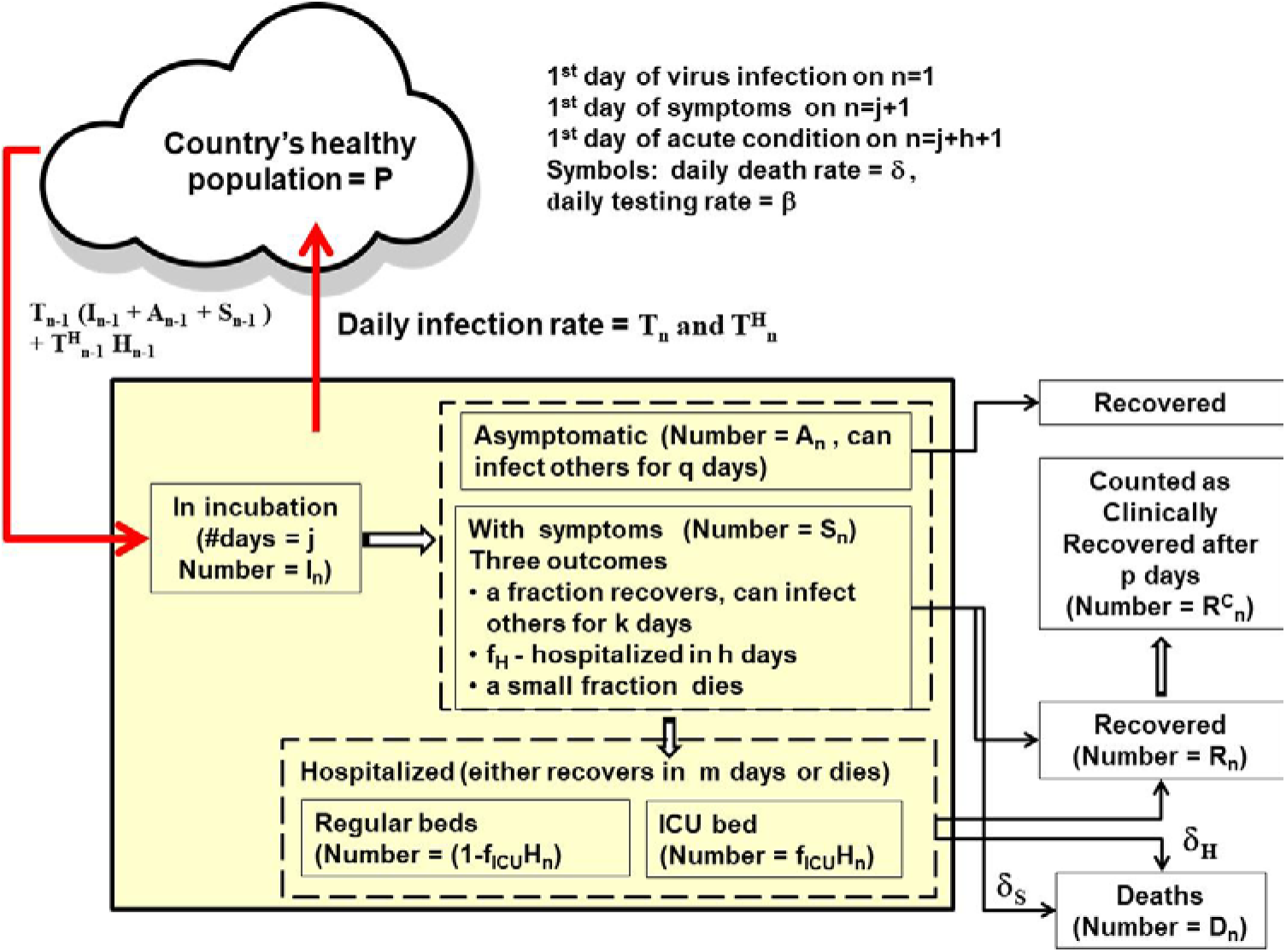
Schematic of the mathematical model for the virus infected cases

On any day n, the total number of people who have been infected with the virus is N_n_, where

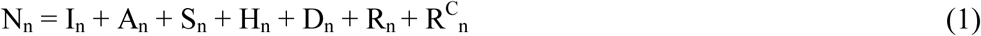

We denote the incubation period for the virus in number of days by j. The model assumes each infected person spends j days in incubation and is capable of infecting others. Let I_n_ be the total population of infected people who are still in the incubation period on day n. Let 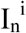 be the population of those infected people in the incubation period on day n who have been infected for i number of days (i ranges from 1 to j).

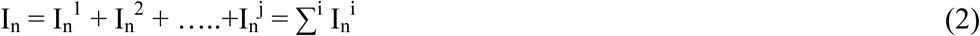

We assume that after the incubation period of j days, each infected person either is asymptomatic or shows symptoms of the disease. Let 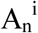 be the population on day n of those asymptomatic people who have been carrying the virus for j+i number of days (1 ≤ i ≤ q) and let A_n_ be the total population of asymptomatic people who are capable of infecting others.

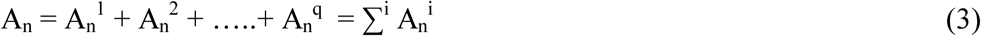

Let 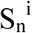 be the population on day n of non-hospitalized patients who have been showing mild symptoms for i number of days (1 ≤ i ≤ k) and let S_n_ be the total population of non-hospitalized patients.

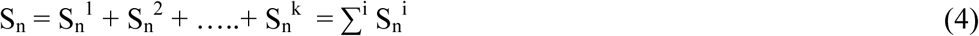

For each person with symptoms, there are three outcomes. A fraction, f_H_ is hospitalized, a small fraction dies at a rate of δ_S_ per person per day, and the remaining fraction recovers. The average time between showing symptoms and hospitalization is h ≤ k. We assume each patient with symptoms is capable of infecting others for a total of j+k number of days.

Let H_n_ be the total population of patients who are hospitalized (in acute condition) on day n. Let 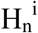 be the population on day n of those infected patients who have been hospitalized for i number of days (i ranges from (1 ≤ i ≤ m).

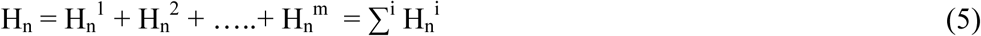

Out of the hospitalized patients, a fraction ficu is admitted to the ICU, the remaining fraction (1-f_ICU_) requires a regular hospital bed. The number of patients in the ICU is

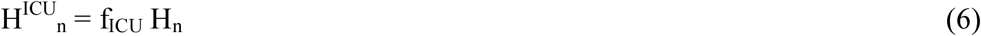

We denote the probability of death among the hospitalized patients to be 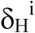 per day and the length of hospital stay to be m number of days. The patients who survive m days in the hospital are recovered but generally needs to wait p more days before being counted as clinically recovered.

The total number of people who are currently infected on day n and who are able to transmit the virus is (I_n_ + A_n_ + S_n_ +H_n_). Out of this population, only (S_n_ + H_n_) number of people show outward signs of the disease. Ironically in the beginning of the outbreak, when (S_n_ + H_n_) is small, the problem is sometimes underestimated without realizing that the disease is spreading exponentially everyday by the entire infected population (I_n_ + A_n_ + S_n_ +H_n_). The ratio (I_n_ + A_n_ + S_n_ +H_n_) / (S_n_ + H_n_) can be very high at the onset of the viral outbreak. The time to take swift, decisive and uniform measures to prevent spreading of the disease is when (S_n_ + H_n_) is small (for example less than 10).

We denote the rate of transmission of the virus from an infected person to a healthy person per day by T_n_. Another way of quantifying the rate of transmission that has been used in the literature is infection reproductive number which for SARS-CoV-2 is reported to be ~ 2.5 [10–11]. Let N_0_ be the number of infected people on the first day. The numbers of infected people on the first two days of the outbreak are:

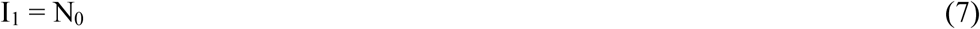

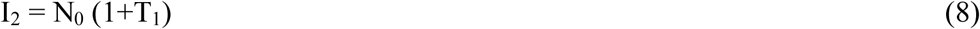

In general, the number of newly infected people on day n is given by

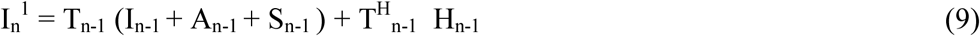

where 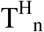 is the rate of transmission from an hospitalized patient to a healthy person. In a well-managed hospital, 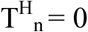. A fraction, f_a_, of the newly infected people is asymptomatic of the disease. An estimate of f_a_ = 30% has been given in [12] which has been used in this model calculations. The number of new patients with mild symptoms on day n is given by

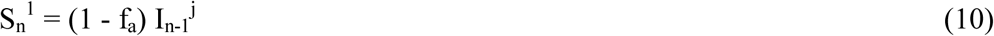

Similarly the number of new asymptomatic patients on day n is given by

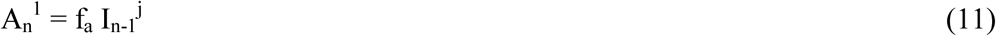

The number of new hospitalized patients on day n is given by

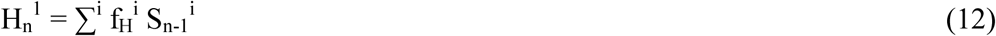

For simplicity, we can assume 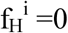 for i ≠ h. The numbers of patients who die and recover from mild symptoms on day n are given by 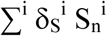 and 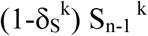 respectively. Similarly the numbers of hospitalized patients who die and recover from acute conditions on day n are given by 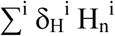 and 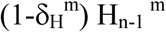 respectively.

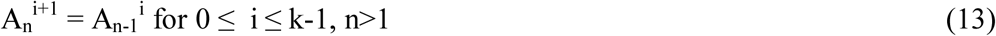

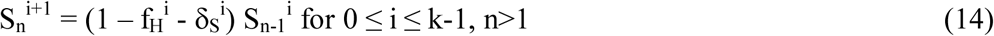

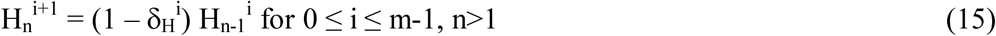

The number of deaths caused by the virus on day n is given by

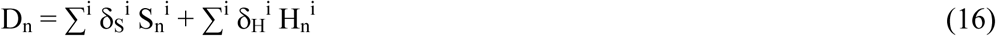

The number of patients who recover from the disease on day n is given by

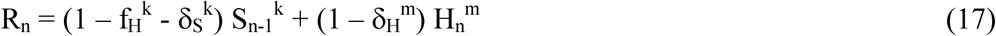

After an infected person first recovers either from the mild symptom state or after being hospitalized, it takes p number of days for her or him to be counted as clinically recovered which may vary from county to county and state to state. The number of new cases of clinically recovered ex-patients on day n is given by

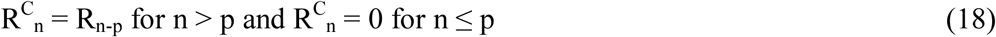

Before the first person showing symptoms of the disease caused by the new virus and shortly thereafter up to day n_1_, the transmission rate T_n_ is assumed to be constant at an initial value T_0_:

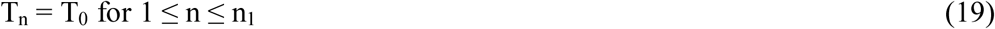

Typically the authorities take steps to prevent the spreading of the disease such as patient isolation and testing, contact tracing and social distancing. As a result T_n_ decreases with time. The model is capable of including actual time dependent transmission data. However at this time such data for the Coronavirus case is not available. A reasonable approximation is exponential decay of the rate of transmission T_n_ from infected persons to the general population. Let G be the number of days it takes for T_n_ - T_f_ to decrease by 63% of T_0_ - T_f_, and let T_f_ be the long term steady state value of T_n_.

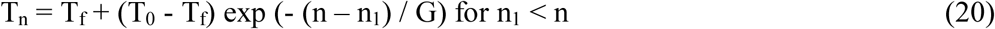

The transmission rate from a hospitalized patient is assumed to be a fraction of T_n_

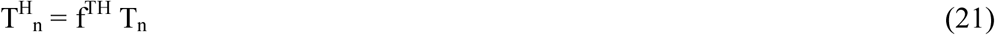

Three parameters n_1_, G and T_f_ in equation (20) were found to be the most significant parameters affecting the number of infected cases and number of deaths from the virus. The data from countries with smaller number of cases could be fit well with smaller n_1_, G and T_f_. In this paper, we will limit the analysis to the first outbreak of the virus in which T_n_ reduces to a final value T_f_. It may also happen that social distancing measures are lifted due to economic or social necessities before the virus is completely extinguished within the population, which would cause T_n_ to rise again from T_f_ which may lead to a recurrence. The recurrence can be modelled in future using the methodology given in this paper.

Let σ_n_ per day per person be the rate of testing of the entire population, P of the country including the infected people without symptoms (I_n_ + A_n_) on any day n. Let β_n_ per day per person be the testing rate of patients with symptoms. Except during the first few days of the virus outbreak, when testing is being developed, β_n_ = 1 in general for people with symptoms.

Cumulative number of cases with symptoms on day n is given by

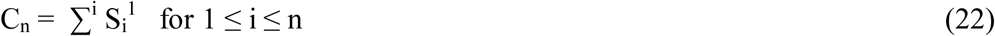

Cumulative number of reported cases which have been positively tested for the virus 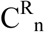 is given by

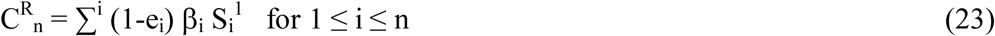

Where e_i_ is the error in reporting the number of patients on i th day. For this paper, we have assumed that the source of data [7–9,13–16] is accurate as of now, and assume e_i_ = 0 for all i. Due to imperfection in the process, often times there is error in reporting the number of cases and also the data is modified from time to time as the accounting procedure changes. The model presented here is flexible and it can incorporate changes in the data set as it is corrected from time to time.

However this model is not capable of predicting suppressed case data.

Cumulative number of deaths on day n is given by

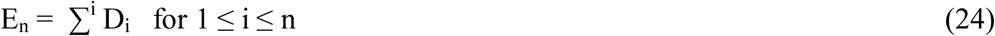

Cumulative number of reported deaths 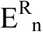 is given by

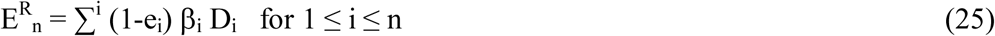

The number of active infected cases which are confirmed by testing and reported on day n can be expressed as:

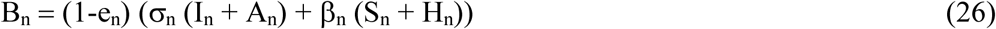

The number of reported hospitalized patients on day n is given by:

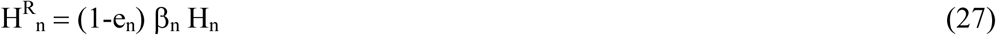

The number of reported hospitalized patients who are in ICU on day n is given by:

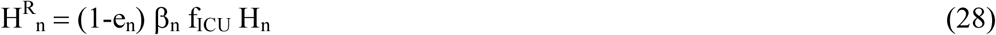

In the model, the testing rates a_n_ and p_n_ are considered time dependent as follows:

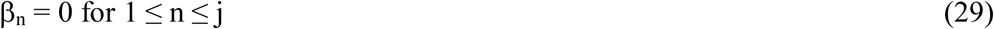

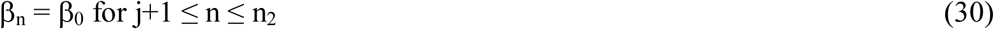

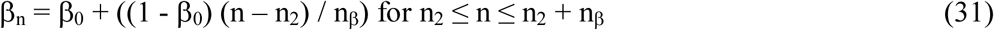

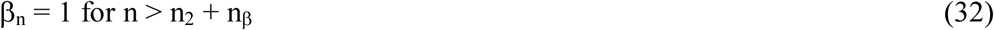

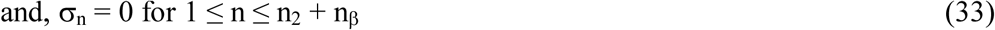

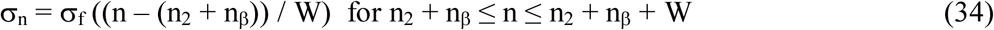

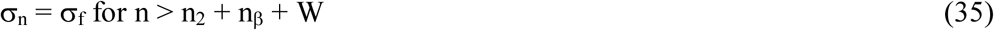

The rationale for choosing different testing rates is as follows. During the first j number of incubation days of the outbreak there is no testing and β_n_ = 0. During the first outbreak of the virus, it takes several days to identify the new virus and to develop an initial test, during which period the testing rate of the patients showing symptoms is low, denoted by β_0_. Let n_β_ be the number of days to ramp up the testing of the patients with symptoms to 100% final testing rate. The testing rate σ_n_ for the general population including those who are infected and not showing symptoms is assumed to be zero until n = n_2_ + n_β_ and let W be number of days it takes to build up the testing capability to test the general population at a final rate of σ_f_. This recognizes that large scale production of testing supplies and development of infrastructure and resources for administering of tests in large numbers take time.

### Model parameters

We test this model to explain the number of Covid-19 cases in a small number of countries at first – Italy, South Korea and New Zealand. These three countries in three continents had very different outcomes of Covid-19 cases and provide a good basis to anchor the model which is designed to be applicable to any country. We collected the data on cumulative total number of cases and deaths for these countries from reliable sources [7–9,13–16] for the period January 1^st^, 2020 to May 8^th^, 2020. We assumed that the aforementioned data sources were reliable and did not verify the accuracy of the data in the datasets. The actual data used in this paper is included in table I in the appendix for the reader to verify. In some cases there is available data on number of recovered cases and active cases. Due to different definitions used in different countries for the criteria used for recovered cases and active cases, we placed much less emphasis on the data on active and recovered cases.

There are twenty one variable parameters in the model description. These are j, k, m, h, p, N_o_, T_o_, T_f_, n_1_, G, β_0_, e, n_2_, n_β_, δ_H_, δ_S_, σ_f_, W, f^TH^, f_H_ and f_ICU_. The strength of this model is that it is based on parameters which can be measured. In the beginning of the virus outbreak, the parameter values are not well known. However in the long run, the parameters values can be extracted from the actual data and the errors in model predictions can be reduced. We start with reasonable assumptions for each parameter and then use the model to fit the published data on total and daily numbers of cases and deaths for each country to iteratively optimize the values of some of the parameters. At the end of iterations, the parameter values that best fit the country data are obtained. With these parameters values, we are in a position

1. to predict the number of cases and required number of hospital beds and
2. to carry out what if analyses on some of the parameters (such as lifting of social distancing requirement)

The initial values of the parameters j, k, m, h, p, f_H_ and f_ICU_ are obtained from data presented in the literature. In this model, we have used j = 5 [18–20], k = 14 [18,20–22], m = 11 [18,20], h = 10 [18,20,22–23], p = 3 [18,21–22], q = 14 [18,20–22], f_H_ = 17% [19–20,24–25], f_ICU_ = 33% [20,24–25]. In addition we used e_i_ = 0 for all i and f_a_ = 30% [12]. The value of p varies widely due to different standards at different places.

It is to be noted that there is daily variation and probability distribution function for each of these model parameters which in principle can be measured. The time and location dependent data on these parameters for Covid-19 in each country is not available yet. When the daily values of these parameters become available on a later date, these can be inserted in the model. For each parameter such as j and m mentioned above there is a range of values. In the model, we used one constant value for each parameter which is very reasonable approximation at this stage of data availability.

We carry out multivariate optimization to fit the data for each country between Jan 1^st^ and May 8^th^ of 2020. This can be considered as the training set. Starting with reasonable initial conditions in the model, iterations are made in the algorithm until the residual errors in fitting the published data during Jan 1^st^ and May 8^th^ 2020 for each country are minimized. At that point the final values of all parameters for this predictive model for each country become available. Once the optimization is completed, the model becomes powerful to yield information such as (1) predicted number of future cases, (2) sensitivity analysis such as the effect of delay in taking initial measures, and (3) recurrence of the disease. We discuss the results in the next section.

## RESULTS AND DISCUSSION

Figure 5 shows the comparison between the Covid-19 case data and the model prediction for Italy. The first reported Covid-19 case in Italy was on Feb 15^th^, 2020. There were 3 total cases on Feb 19 and the first death was on Feb 21. The model fitted the Italy Covid-19 case data with Feb 22 start date for transmission factor decrease when the number of cases was 79 and a transmission decay rate of 1/G, where G= 14. The model fits the data very well. The model predicts the number of cases to rise to between 232,000 and 239,000 and the number of deaths to rise to between 34,000 and 35,000 on June 30^th^, 2020.

**Figure 5.**
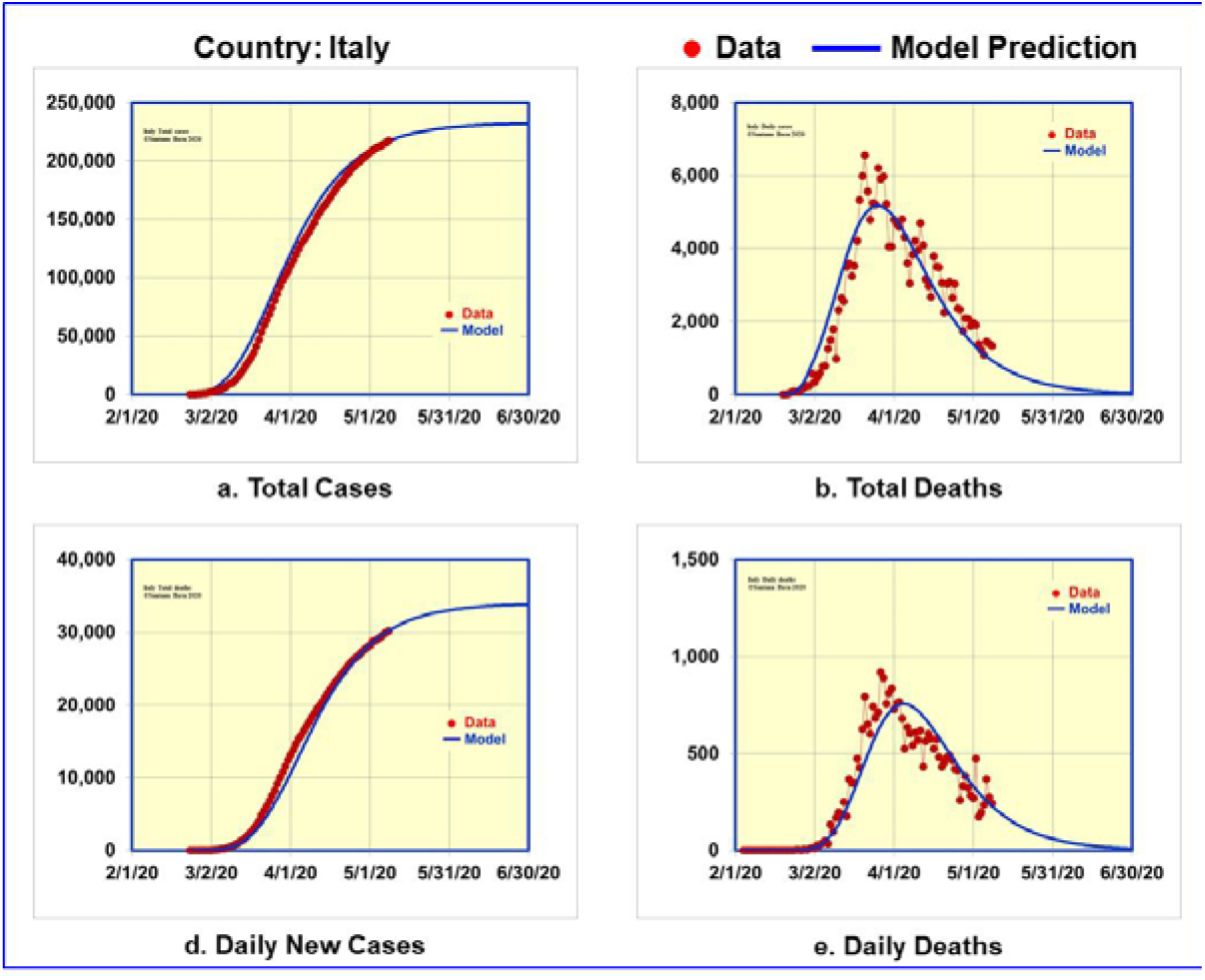
Data and model predictions for Covid-19 cases in Italy

Figure 6 shows the comparison of the actual data and the model predictions for South Korea which had a factor of twenty less number of cases than Italy. The daily case data for South Korea has been available since Feb 15^th^. There were 28 total cases on Feb 15^th^ and the first Covid-19 death was on Feb 20^th^. The model fitted the South Korea Covid-19 data with Feb 18^th^ start date for transmission factor decrease when the number of cases was 31 and G= 6.

**Figure 6.**
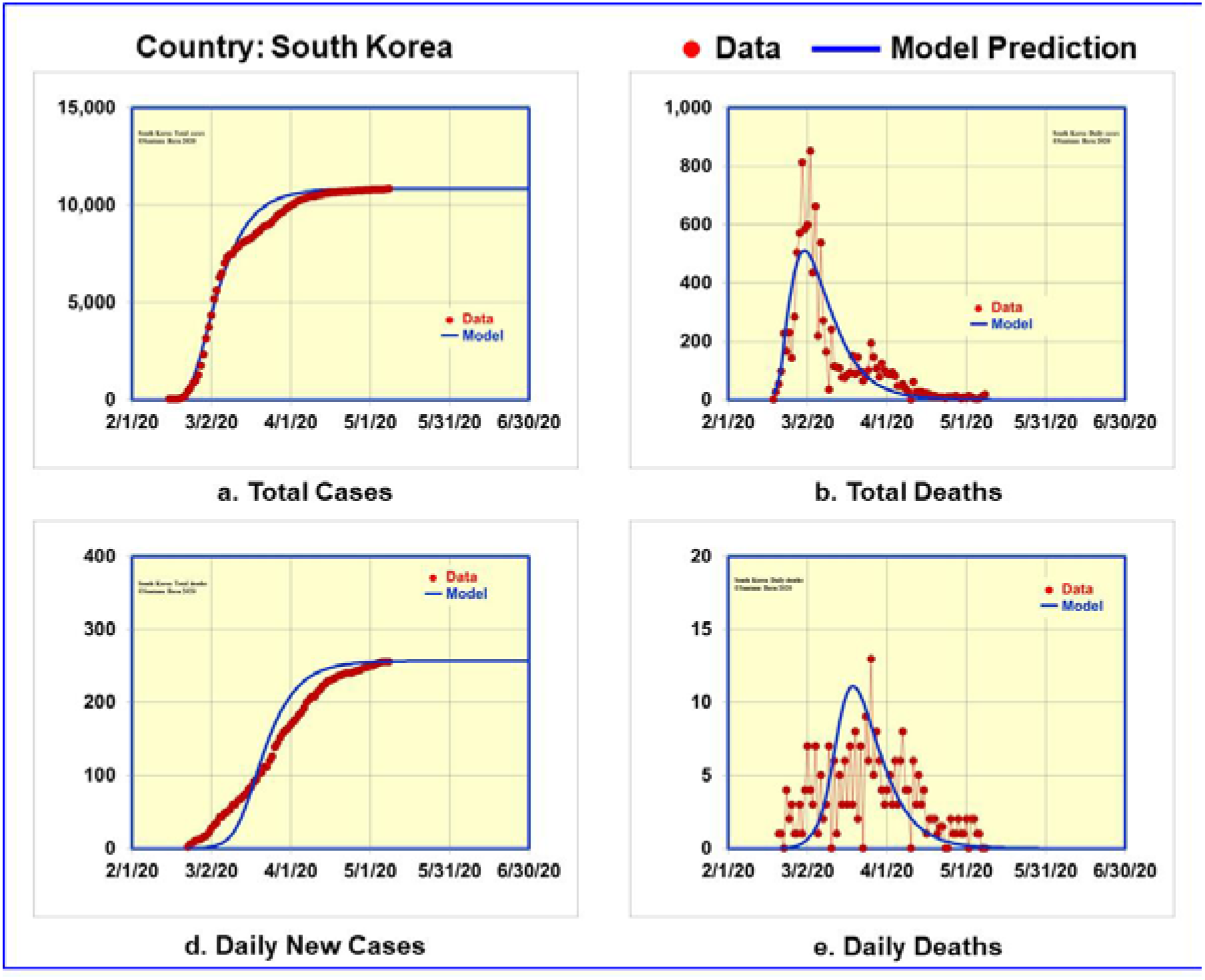
Data and model predictions for Covid-19 cases in South Korea

Unless there is a recurrence, the model does not predict any significant rise in number of cases or deaths by June 30^th^, 2020. Comparing model parameters for Italy and South Korea, we found that the number of cases were lower when T_n_ started decreasing and G was smaller for South Korea.

Figure 7 shows the comparison of the actual data and the model predictions for New Zealand which had a factor of seven less number of cases than South Korea. The daily case data for New Zealand became available since Mar 6^th^ on which date there were 4 total cases. The first Covid-19 death was on Mar 28^th^. The model fitted the New Zealand Covid-19 data using the start date of reduction in T_n_ when the total number of reported cases was only 8, and the transmission factor to decay in G= 10 days. Unless there is a recurrence, the model does not predict any significant rise in number of cases or deaths in New Zealand by June 30^th^, 2020.

**Figure 7.**
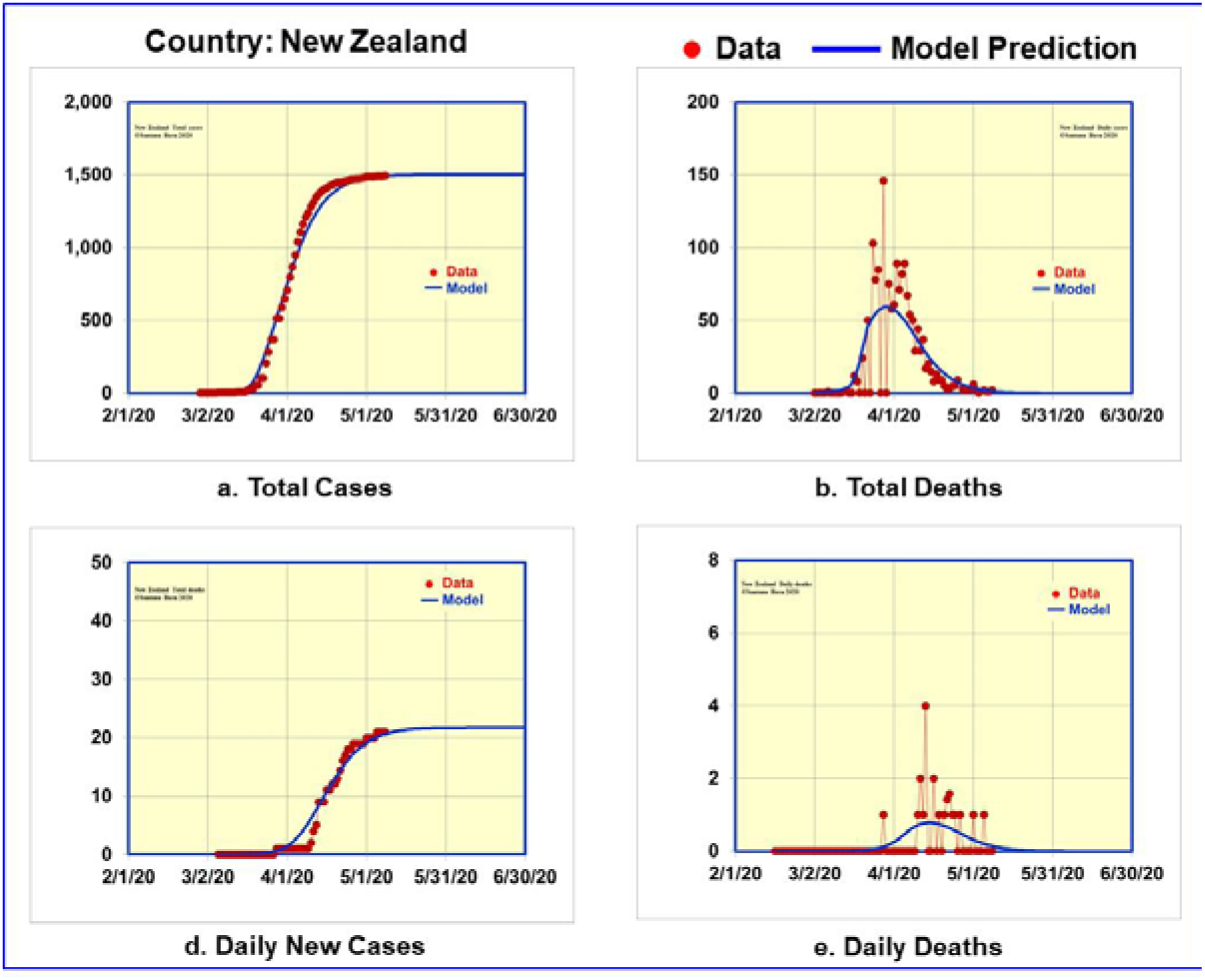
Data and model predictions for Covid-19 cases in New Zealand

Comparing data from Italy, South Korea and New Zealand, the model makes it obvious that when a country takes timely, swift and uniform virus containment measures at the point when the total number of cases is small, the number of infected cases and deaths are small. It is very similar to containment strategy for wildfire: sooner to detect, sooner to start containment, the less the spread and the resulting destruction. Even a day of waiting makes a big difference in the case of Coronavirus. In the model, transmission factors started reducing for Italy, South Korea and New Zealand when the total cases were 79, 31 and 8 respectively.

In conclusion, we have developed a mathematical model that provides insight into the Covid-19 viral outbreak cases in any country. The model fits the data very well for Italy, South Korea, New Zealand and California with different levels of cases and casualties. We also presented predictions from the model for hospital bed requirements six weeks in advance which can be very valuable for providing medical services to reduce casualties. Since conditions change daily which causes the model parameters to change, we do not predict more than six weeks in advance. The model is transparent and is based on parameters which are measurable. This allows the model calculations to be updated as the actual data of the parameters become available. Overall the model makes the hopeful conclusion that Covid-19 can be effectively contained by taking timely social distancing and patient isolation measures before ultimately a vaccine becomes available.

The author wishes to acknowledge valuable suggestion from Professor Nigam Shah of Stanford University to look into the hospitalization needs.

## Data Availability

The data is available in the paper.

## Additional Information

Correspondence and requests for materials should be addressed to the author.

## APPENDIX

**Table I.** Covid-19 case data for South Korea, Italy, New Zealand and California (Jan 1- May 8, 2020)

| Date | South Korea |  | Italy |  | New Zealand |  | California |  |  |  |
| --- | --- | --- | --- | --- | --- | --- | --- | --- | --- | --- |
|  | Total cases | Total deaths | Total cases | Total deaths | Total cases | Total deaths | Total cases | Total deaths | Hospital patients on day | ICU patients on day |
| 14-Feb-20 |  |  |  |  |  |  |  |  |  |  |
| 15-Feb-20 | 28 |  | 3 |  |  |  |  |  |  |  |
| 16-Feb-20 | 29 |  | 3 |  |  |  |  |  |  |  |
| 17-Feb-20 | 30 |  | 3 |  |  |  |  |  |  |  |
| 18-Feb-20 | 31 |  | 3 |  |  |  |  |  |  |  |
| 19-Feb-20 | 58 |  | 3 |  |  |  |  |  |  |  |
| 20-Feb-20 | 111 | 1 | 4 |  |  |  |  |  |  |  |
| 21-Feb-20 | 209 | 2 | 21 | 1 |  |  |  |  |  |  |
| 22-Feb-20 | 436 | 2 | 79 | 2 |  |  |  |  |  |  |
| 23-Feb-20 | 602 | 6 | 157 | 3 |  |  |  |  |  |  |
| 24-Feb-20 | 833 | 8 | 229 | 7 |  |  |  |  |  |  |
| 25-Feb-20 | 977 | 11 | 323 | 11 |  |  |  |  |  |  |
| 26-Feb-20 | 1261 | 12 | 470 | 12 |  |  |  |  |  |  |
| 27-Feb-20 | 1766 | 13 | 655 | 17 |  |  |  |  |  |  |
| 28-Feb-20 | 2337 | 16 | 889 | 21 | 1 |  |  |  |  |  |
| 29-Feb-20 | 3150 | 17 | 1128 | 29 | 1 |  |  |  |  |  |
| 01-Mar-20 | 3736 | 21 | 1701 | 41 | 1 |  |  |  |  |  |
| 02-Mar-20 | 4335 | 28 | 2036 | 52 | 1 |  |  |  |  |  |
| 03-Mar-20 | 5186 | 32 | 2502 | 79 | 2 |  |  |  |  |  |
| 04-Mar-20 | 5621 | 35 | 3089 | 107 | 3 |  | 53 | 1 |  |  |
| 05-Mar-20 | 6284 | 42 | 3858 | 148 | 4 |  | 60 | 1 |  |  |
| 06-Mar-20 | 6503 | 43 | 4636 | 197 | 4 |  | 69 | 1 |  |  |
| 07-Mar-20 | 7041 | 48 | 5883 | 233 | 5 |  | 88 | 1 |  |  |
| 08-Mar-20 | 7313 | 50 | 7375 | 366 | 5 |  | 114 | 1 |  |  |
| 09-Mar-20 | 7478 | 53 | 9172 | 463 | 5 |  | 133 | 1 |  |  |
| 10-Mar-20 | 7513 | 60 | 10149 | 631 | 5 |  | 157 | 2 |  |  |
| 11-Mar-20 | 7755 | 60 | 12462 | 827 | 5 |  | 177 | 3 |  |  |
| 12-Mar-20 | 7869 | 66 | 15113 | 1016 | 5 |  | 198 | 4 |  |  |
| 13-Mar-20 | 7979 | 67 | 17660 | 1266 | 6 |  | 247 | 5 |  |  |
| 14-Mar-20 | 8086 | 72 | 21157 | 1441 | 8 |  | 335 | 6 |  |  |
| 15-Mar-20 | 8162 | 75 | 24747 | 1809 | 8 |  | 392 | 6 |  |  |
| 16-Mar-20 | 8236 | 81 | 27980 | 2158 | 8 |  | 472 | 11 |  |  |
| 17-Mar-20 | 8320 | 84 | 31506 | 2503 | 20 |  | 598 | 13 |  |  |
| 18-Mar-20 | 8413 | 91 | 35713 | 2978 | 28 |  | 675 | 16 |  |  |
| 19-Mar-20 | 8565 | 94 | 41035 | 3405 | 28 |  | 1006 | 19 |  |  |
| 20-Mar-20 | 8652 | 102 | 47021 | 4032 | 52 |  | 1224 | 23 |  |  |
| 21-Mar-20 | 8799 | 104 | 53578 | 4825 | 52 |  | 1468 | 27 |  |  |
| 22-Mar-20 | 8897 | 111 | 59138 | 5476 | 102 |  | 1733 | 27 |  |  |
| 23-Mar-20 | 8961 | 111 | 63927 | 6077 | 102 |  | 2102 | 40 |  |  |
| 24-Mar-20 | 9037 | 120 | 69176 | 6820 | 205 |  | 2535 | 53 |  |  |
| 25-Mar-20 | 9137 | 126 | 74386 | 7503 | 283 |  | 3006 | 65 |  |  |
| 26-Mar-20 | 9332 | 139 | 80589 | 8215 | 368 |  | 3801 | 78 | 746 | 200 |

Table I (contd.). Covid-19 case data for South Korea, Italy, New Zealand and California (Jan 1- May 8, 2020)
| Date | South Korea |  | Italy |  | New Zealand |  | California |  |  |  |
| --- | --- | --- | --- | --- | --- | --- | --- | --- | --- | --- |
|  | Total cases | Total deaths | Total cases | Total deaths | Total cases | Total deaths | Total cases | Total deaths | Hospital patients on day | ICU patients on day |
| 27-Mar-20 | 9478 | 144 | 86498 | 9134 | 368 |  | 4643 | 101 |  |  |
| 28-Mar-20 | 9583 | 152 | 92472 | 10023 | 514 | 1 | 4643 | 101 |  |  |
| 29-Mar-20 | 9661 | 158 | 97689 | 10779 | 514 | 1 | 5763 | 135 | 1307 | 545 |
| 30-Mar-20 | 9786 | 162 | 101739 | 11591 | 589 | 1 | 6932 | 150 | 1629 | 665 |
| 31-Mar-20 | 9887 | 165 | 105792 | 12428 | 647 | 1 | 8155 | 171 | 1855 | 774 |
| 01-Apr-20 | 9976 | 169 | 110574 | 13155 | 708 | 1 | 9191 | 203 | 1922 | 816 |
| 02-Apr-20 | 10062 | 174 | 115242 | 13915 | 797 | 1 | 10701 | 237 | 2188 | 901 |
| 03-Apr-20 | 10156 | 177 | 119827 | 14681 | 868 | 1 | 12026 | 276 | 2300 | 1008 |
| 04-Apr-20 | 10237 | 183 | 124632 | 15362 | 950 | 1 | 13438 | 319 | 2398 | 1040 |
| 05-Apr-20 | 10284 | 186 | 128948 | 15887 | 1039 | 1 | 14336 | 343 | 2509 | 1085 |
| 06-Apr-20 | 10331 | 192 | 132547 | 16523 | 1106 | 1 | 15865 | 374 | 2611 | 1108 |
| 07-Apr-20 | 10384 | 200 | 135586 | 17127 | 1160 | 1 | 16957 | 442 | 2714 | 1154 |
| 08-Apr-20 | 10423 | 204 | 139422 | 17669 | 1210 | 1 | 18309 | 492 | 2825 | 1132 |
| 09-Apr-20 | 10450 | 208 | 143626 | 18279 | 1239 | 1 | 19472 | 541 | 2897 | 1145 |
| 10-Apr-20 | 10450 | 208 | 147577 | 18849 | 1283 | 2 | 20615 | 609 | 2846 | 1145 |
| 11-Apr-20 | 10512 | 214 | 152271 | 19468 | 1312 | 4 | 21794 | 651 | 2917 | 1145 |
| 12-Apr-20 | 10537 | 217 | 156363 | 19899 | 1349 | 5 | 22348 | 687 | 3015 | 1178 |
| 13-Apr-20 | 10564 | 222 | 159516 | 20465 | 1366 | 9 | 23338 | 758 | 3124 | 1177 |
| 14-Apr-20 | 10591 | 225 | 162488 | 21067 | 1386 | 9 | 24424 | 821 | 3171 | 1175 |
| 15-Apr-20 | 10613 | 229 | 165155 | 21645 | 1401 | 9 | 26182 | 890 | 3141 | 1191 |
| 16-Apr-20 | 10635 | 230 | 168941 | 22170 | 1409 | 11 | 27528 | 985 | 3180 | 1175 |
| 17-Apr-20 | 10653 | 232 | 172434 | 22745 | 1422 | 11 | 28963 | 1072 | 3221 | 1174 |
| 18-Apr-20 | 10661 | 234 | 175925 | 23227 | 1431 | 12 | 30333 | 1166 | 3196 | 1163 |
| 19-Apr-20 | 10674 | 236 | 178972 | 23660 | 1440 | 12 | 30978 | 1208 | 3257 | 1196 |
| 20-Apr-20 | 10683 | 237 | 181228 | 24114 | 1445 | 13 | 33261 | 1268 | 3365 | 1241 |
| 21-Apr-20 | 10692 | 238 | 184252 | 24595 | 1448 | 14 | 35396 | 1354 | 3357 | 1219 |
| 22-Apr-20 | 10702 | 240 | 187327 | 25085 | 1451 | 16 | 37369 | 1469 | 3343 | 1204 |
| 23-Apr-20 | 10708 | 240 | 189973 | 25549 | 1456 | 17 | 39254 | 1562 | 3344 | 1216 |
| 24-Apr-20 | 10718 | 240 | 192994 | 25969 | 1461 | 18 | 41137 | 1651 | 3343 | 1198 |
| 25-Apr-20 | 10728 | 242 | 195351 | 26384 | 1470 | 18 | 42164 | 1710 | 3324 | 1184 |
| 26-Apr-20 | 10738 | 243 | 197675 | 26644 | 1469 | 19 | 43464 | 1755 | 3343 | 1185 |
| 27-Apr-20 | 10752 | 244 | 199414 | 26977 | 1472 | 19 | 45031 | 1809 | 3455 | 1181 |
| 28-Apr-20 | 10761 | 246 | 201505 | 27359 | 1474 | 19 | 46500 | 1887 | 3495 | 1186 |
| 29-Apr-20 | 10765 | 247 | 203591 | 27682 | 1476 | 19 | 48917 | 1982 | 3497 | 1192 |
| 30-Apr-20 | 10774 | 248 | 205463 | 27967 | 1479 | 19 | 50442 | 2073 | 3428 | 1192 |
| 01-May-20 | 10780 | 250 | 207428 | 28236 | 1485 | 20 | 52197 | 2171 | 3423 | 1179 |
| 02-May-20 | 10793 | 250 | 209328 | 28710 | 1487 | 20 | 53616 | 2215 | 3423 | 1169 |
| 03-May-20 | 10801 | 252 | 210717 | 28884 | 1487 | 20 | 54,937 | 2,254 | 3284 | 1179 |
| 04-May-20 | 10804 | 254 | 211938 | 29079 | 1486 | 20 | 56,212 | 2,317 | 3369 | 1157 |
| 05-May-20 | 10806 | 255 | 213013 | 29315 | 1488 | 21 | 58,815 | 2,412 | 3334 | 1140 |
| 06-May-20 | 10810 | 256 | 214457 | 29684 | 1489 | 21 | 60,614 | 2,504 | 3319 | 1125 |
| 07-May-20 | 10822 | 256 | 215858 | 29958 | 1490 | 21 | 62,512 | 2,585 | 3250 | 1118 |
| 08-May-20 | 10840 | 256 | 217185 | 30201 | 1492 | 21 | 64,561 | 2,678 | 3282 | 1115 |

## Notes

### Competing Interest Statement

The authors have declared no competing interest.

### Funding Statement

This work was funded by Sparkle Optics

### Author Declarations

This is a modelling paper approved by Sparkle Optics

